# BNT162b2 vaccination induces SARS-CoV-2 specific antibody secretion into human milk with minimal transfer of vaccine mRNA

**DOI:** 10.1101/2021.04.27.21256151

**Authors:** Jia Ming Low, Yue Gu, Melissa Shu Feng Ng, Zubair Amin, Le Ye Lee, Yvonne Peng Mei Ng, Bhuvaneshwari D/O Shunmuganathan, Yuxi Niu, Rashi Gupta, Paul Anantharajah Tambyah, Paul A MacAry, Liang Wei Wang, Youjia Zhong

## Abstract

**Importance:** To examine the impact of SARS-CoV-2 vaccination of lactating mothers on human milk

**Objective:** (1) To quantify SARS-CoV-2-specific immunoglobulin A (IgA) and immunoglobulin G (IgG) in human milk of lactating mothers who received the BNT162b2 vaccine, with reference to a cohort convalescent from antenatal COVID-19, and healthy lactating mothers. (2) To detect and quantify vaccine mRNA in human milk after BNT162b2 vaccination.

**Design:** Gestational Immunity For Transfer 2 (GIFT-2) is a prospective cohort study of lactating mothers who were due to receive two doses of BNT162b2 vaccine, recruited between 5th February 2021 and 9th February 2021.

**Setting:** Lactating healthcare workers living in Singapore

**Participants:** Convenience sample of ten lactating healthcare workers. Human milk samples were collected at four time points: pre-vaccination, 1-3 days after dose one, 7-10 days after dose one, and 3-7 days after dose two of the BNT162b2 vaccine.

**Exposure:** Two doses of the BNT162b2 vaccine 21 days apart.

**Main Outcome and Measure:** (i) SARS-CoV-2-specific IgA and IgG in human milk of lactating mothers who received BNT162b2 vaccine, (ii) Detection and quantification of vaccine mRNA in human milk after BNT162b2 vaccination.

**Results:** Ten lactating healthcare workers aged 32.5 years (range 29 – 42) were recruited, with 40 human milk samples collected and analysed. SARS-CoV-2-specific IgA was predominant in human milk of lactating mothers who received BNT162b2 vaccine. The sharpest rise in antibody production was 3 −7 days after dose two of the BNT162b2 vaccine, with medians of 1110 picomolar of anti-SARS-CoV-2 spike and 374 picomolar of anti-Receptor Binding Domain IgA. Vaccine mRNA was detected only on rare occasions, at a maximum concentration of 2 ng/mL.

**Conclusions and Relevance:** In this cohort of ten lactating mothers following BNT162b2 vaccination, nine (90%) produced SARS-CoV-2 IgA, and ten (100%) produced IgG in human milk with minimal amounts of vaccine mRNA. Lactating individuals should continue breastfeeding in an uninterrupted manner after receiving mRNA vaccination for SARS-CoV-2.

**Trial Registration:** Registered at clinicaltrials.gov (NCT04802278).

**Key Points:** *Question:* Does BNT162b2 (i) induce the production and secretion of SARS-CoV-2 specific antibodies into human milk, and/or (ii) get secreted into human milk?

*Findings:* In this cohort that included ten lactating healthcare workers following BNT162b2 vaccination, 90% produced SARS-CoV-2 immunoglobulin A, and 100% produced immunoglobulin G in human milk, with minimal amounts of vaccine mRNA transfer.

*Meaning:* Lactating individuals should continue breastfeeding in an uninterrupted manner after receiving SARS-CoV-2 mRNA vaccination.

## Background

Lactating mothers who have recovered from respiratory virus infections produce antibodies in human milk that can neutralize offending viruses *in vivo*.^1-3^ The production of protective antibodies in human milk after influenza vaccination in lactating mothers confers local mucosal immunity to infants and has informed antenatal vaccination programs for diseases such as influenza and tetanus in lactating mothers.^4,5^

However, similar evidence for Coronavirus Disease-19 (COVID-19) messenger ribonucleic acid (mRNA) vaccines is scarce.^6^ As a result, vaccine advisories from various health authorities have been cautious in recommending vaccinations for lactating mothers. The American College of Obstetricians and Gynecologists (ACOG) and the Royal College of Obstetricians & Gynaecologists (RCOG, UK) both state that COVID-19 vaccines may be offered to lactating individuals, while acknowledging that adequate safety data is not available.^7,8^ In countries such as Singapore, mothers have been advised to express and discard human milk for up to seven days after vaccination.^9^ However, such measures may disrupt mother-child bonding and may result in premature cessation of breastfeeding.^10^

There is emerging evidence that SARS-CoV-2 specific antibodies are detectable in human milk post vaccination;^11-14^ however, the amount of immunoglobulin G (IgG) and immunoglobulin A (IgA) have not been clearly quantified. It is also unknown whether vaccine components such as mRNA are transferred in human milk; preliminary studies purportedly detect no vaccine mRNA in human milk.^15^

Our aims are (1) to quantify SARS-CoV-2-specific IgA and IgG in human milk of lactating mothers who received COVID-19 mRNA vaccine, with reference to a cohort convalescent from antenatal COVID-19 as well as a control cohort of healthy lactating women) and (2) to detect and quantify vaccine mRNA in human milk after vaccination.

We hypothesize that BNT162b2, an mRNA vaccine encoding the immunogenic SARS-CoV-2 spike protein, has minimal leakage into human milk after vaccination, and induces the production and secretion of spike- and receptor-binding domain (RBD)-specific IgA and IgG into human milk.

## Methods

We conducted a prospective cohort study of a convenience sample of lactating healthcare workers living in Singapore, who were due to receive two doses of the BNT162b2 (Pfizer/BioNtech) vaccine. The study was approved by the Institutional Review Board (Gestational Immunity For Transfer GIFT-2: DSRB Reference Number: 2021/00095) and the study protocol was registered at clinicaltrials.gov (NCT04802278).^16^ Participants were recruited between 5th February 2021 and 9th February 2021 and were invited to participate via advertisements and social media. Exclusion criteria were that of no prior known exposure to COVID-19, and any autoimmune disease, current or recent infections, cancer, and any current or recent immunomodulatory medication. Demographic details and clinical outcomes of lactating mothers and their infants were determined through a structured questionnaire up to 28 days following mother’s second dose of vaccine.

Human milk samples from the “vaccinated cohort” were collected at four time points: pre-vaccination (T1),1-3 days after dose one (T2), 7-10 days after dose one (T3), and 3-7 days after dose two of COVID-19 mRNA vaccine (T4). All samples were collected at home using electric breast pumps or hand expressed into sterile plastic containers, stored and transported via courier immediately at −18°C to the laboratory. In the laboratory, the samples were stored at −80°C until analysis.

Human milk samples at 1 month postpartum from convalescent and healthy mothers were also analysed for reference. The “convalescent cohort” were six women who had COVID-19 in pregnancy confirmed with real-time polymerase-chain-reaction (RT-PCR) assay, and then recovered as defined by resolution of clinical symptoms and with two negative RT-PCR assays 24 hours apart. Nine healthy unrelated mothers without COVID-19 infection or vaccination served as the “healthy cohort”.

Demographic details and clinical outcomes of lactating mothers and their infants were determined up to 28 days through a structured questionnaire after ingestion of post-vaccination human milk.

Assays performed on milk samples were: (1) quantitative ELISA for spike- and RBD-specific IgA and IgG against SARS-CoV-2 antigens, and (2) sensitive quantitation of BNT162b2 mRNA.

Human milk was first tested for presence of IgA in all four time points (i.e. T1-T4), whereas T1 and T4 were tested for the presence of IgG, as IgA levels were only seen to rise at T4.

IgA and IgG against SARS-CoV-2 spike and RBD were titrated using a quantitative ELISA. A human monoclonal antibody specific for SARS-CoV-2 was recombinantly engineered and expressed as human IgG1 and/or IgA1. This was employed as the reagent for quantitation (details available in eMethods in the Supplement).

Total ribonucleic acid (RNA) from whole human milk was extracted for BNT162b2 mRNA detection. Phenol-chloroform extraction was utilised to increase mRNA yield and used as input for polyA-primed reverse transcription. A standard curve was constructed using BNT162b2 vaccine spiked into healthy SARS-CoV-2 negative human milk and then extracted in the aforementioned manner to allow us to quantify the mRNA detected. This also served as the positive control. Detection of BNT162b2 mRNA was then performed using probe-based quantitative PCR (qPCR; details available in eMethods in the Supplement).

Clinical characteristics of the cohorts were summarised using GraphPad Prism 8. Shapiro-Wilk test of normality was used; if data was normal, mean and standard deviations were presented. The rest of the statistical analyses were done in R (4.0.2). Two groups were compared with Mann-Whitney U test (two tailed). For multiple comparisons, the PMCMRplus package (PMCMRplus_1.9.0) was used to perform Kruskal-Wallis test with Dunn’s nonparametric all-pairs comparison post-test. p<0.05 level of confidence was accepted for statistical significance. Details of sample sizes and analyses performed specific to each figure are in all figure legends. All box and whiskers plots show median (center line), interquartile range (box), and 10th and 90th percentiles (whiskers). All data points are plotted with the boxplots. All line plots show the median, and error bars show the first and third quartiles. The study is reported in accordance to the STROBE reporting guidelines for cohort studies.^17^

## Results

### Maternal and infant characteristics

Ten lactating healthcare workers were recruited; all received two doses of the BNT162b2 (Pfizer/BioNtech) vaccine, with the second dose given on day 21. The mothers were of a mean age of 33.5 [standard deviation, SD 3.3] years of age, eight (80%) were Singaporean Chinese and two (20%) were Singaporean Malay. At recruitment, mothers were a mean of 9.8 [SD 4.0] months post-partum. (Table 1) Samples for T1-T4 were available for all subjects so 40 samples were collected and analysed in total. All infants were born full term and healthy. No mother or infant experienced any serious adverse event during the 28-day study period. None of the mothers had mastitis after vaccination. Nine infants were breastfed within 72 hours after their mothers were vaccinated; one infant was not fed breast milk within 72 hours after vaccination. None of these nine developed any fever, rash, vomiting, diarrhea, cough or rhinorrhoea, up to 28 days after ingestion of post-vaccination human milk.

**Table 1:**
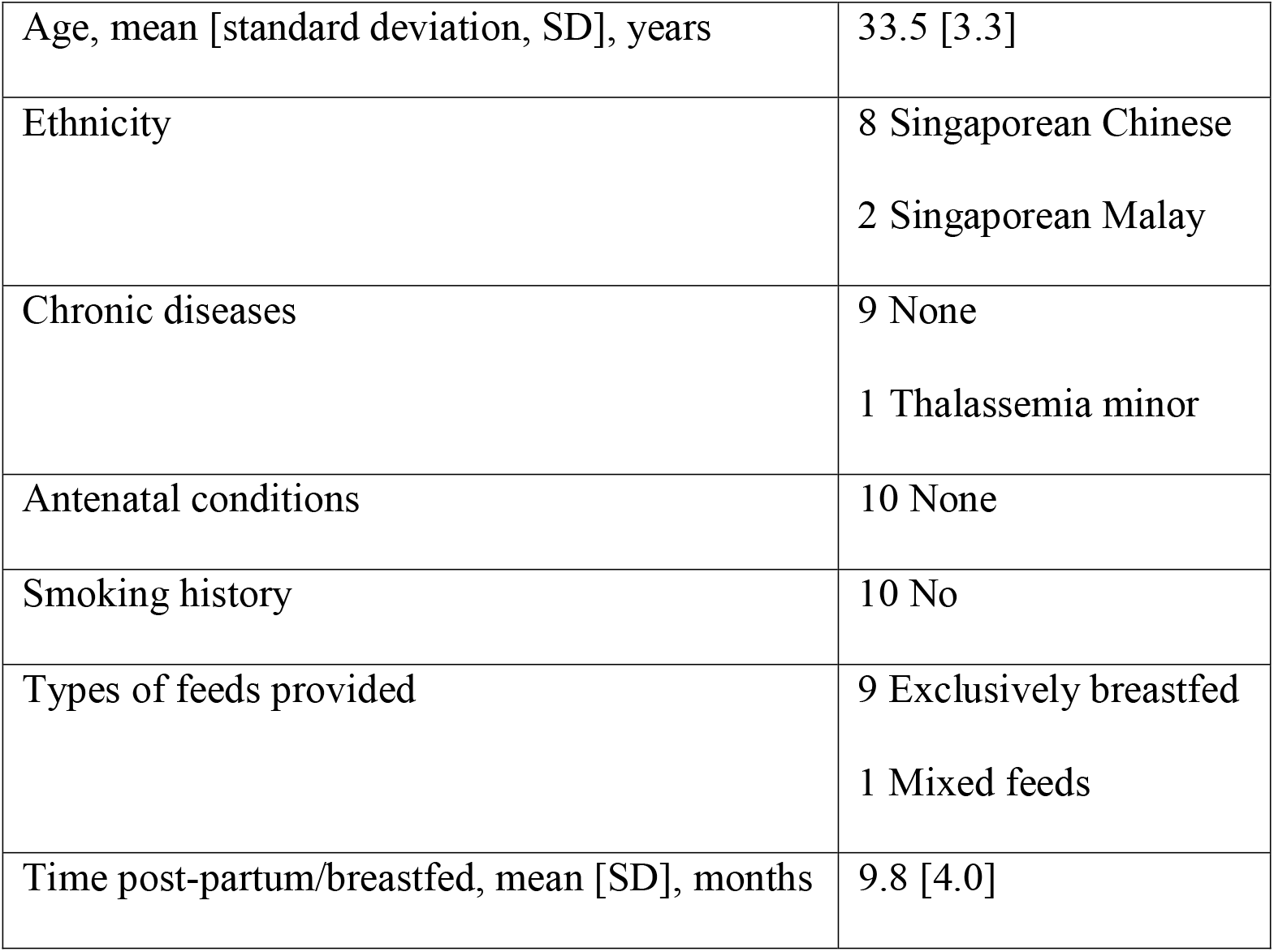
Characteristics of vaccinated mothers.

Women in the reference cohort had COVID-19 with a mean of 136 [SD 80] days before delivery; thus, human milk collected one month postpartum was about 5.5 months from the point of initial antigenic exposure. Subjects in the convalescent and healthy women did not receive SARS-CoV-2 vaccination during the study. Clinical details of the convalescent and healthy women in the reference cohort are available in the eTables 1 and 2 in the Supplement.

### Levels of SARS-CoV-2-specific IgA and IgG in in human milk of lactating mothers

A human monoclonal antibody binding to SARS-CoV-2 was recombinantly engineered and expressed as human IgG and human IgA. This resulted in the production of monoclonal antibodies of different isotypes with the same antigen-binding ability, which were utilized as reference reagents for construction of standard curves for the quantitative ELISA (eFigure1 in the Supplement). Human milk samples were evaluated for IgA binding reactivity against SARS-CoV-2 spike and RBD (Figure 1; eFigure 2 in the Supplement).

**Figure 1.**
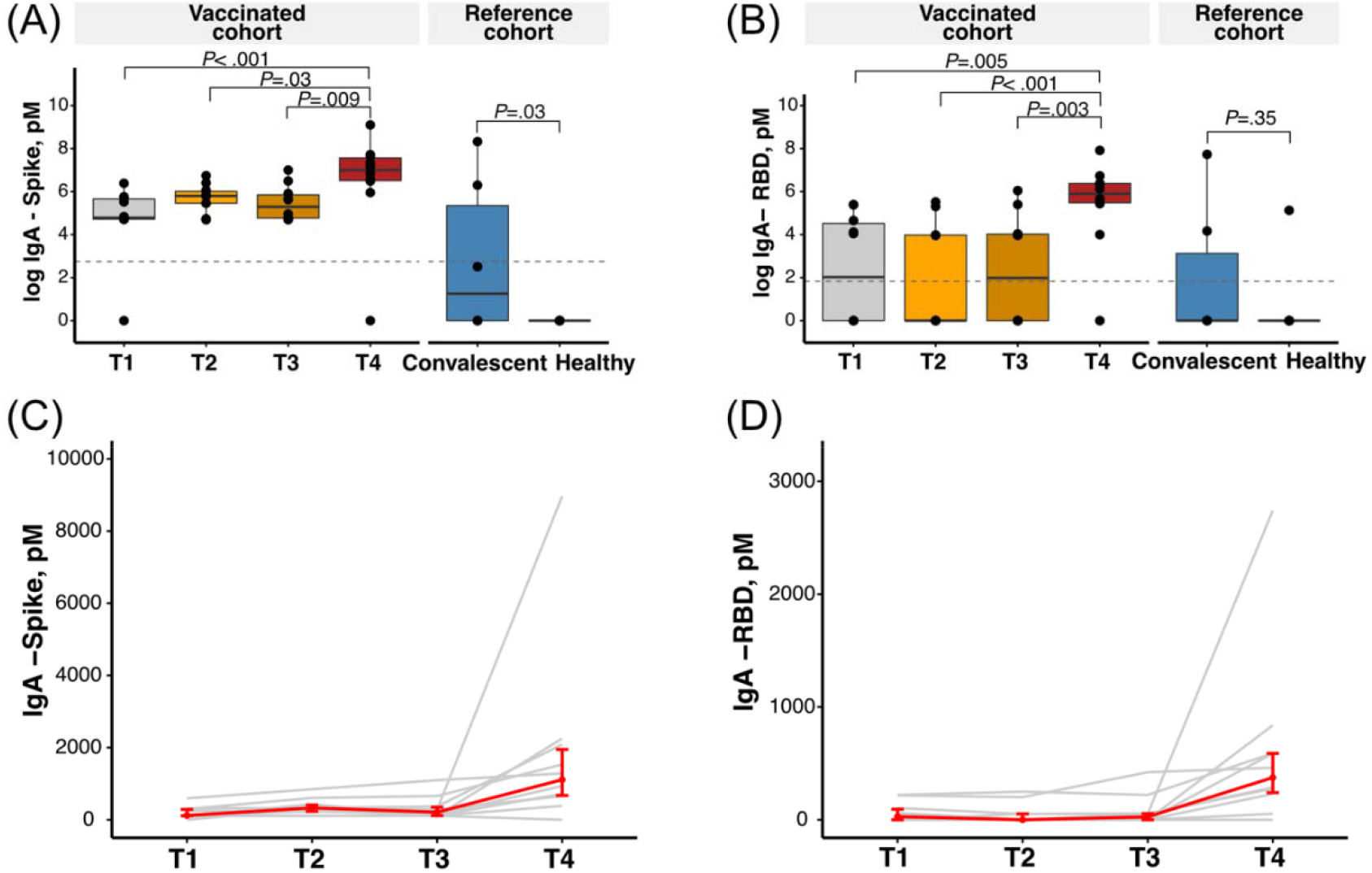
IgA antibodies in human milk samples. (A) & (B): Concentration [pM^a^] of IgA antibodies against spike (A) and RBD (B) in human milk for vaccinated mothers (n =10) across all time points; convalescent (n = 6) and healthy mothers (n = 9) used as reference cohort. Grey dotted lines reflect the limit of assay detection. Statistics were calculated with Kruskal-Wallis test and Dunn’s post-test. (C) & (D): Increase in IgA antibodies against spike (C) and RBD (D) over time. Each line in grey represents data from one individual and the median ± IQR is represented in red. Abbreviations: RBD, receptor-binding domain; IgA, immunoglobulin A; pM, picomolar; IQR, interquartile range; T1, pre-vaccination; T2, 1-3 days after dose one of BNT162b2 vaccine; T3, 7-10 days after dose one of BNT162b2 vaccine; T4, 3-7 days after dose two of BNT162b2 vaccine. ^a^ SI conversion factors: To convert concentration from pM to M, multiply values by 10^12^.

**Figure 2.**
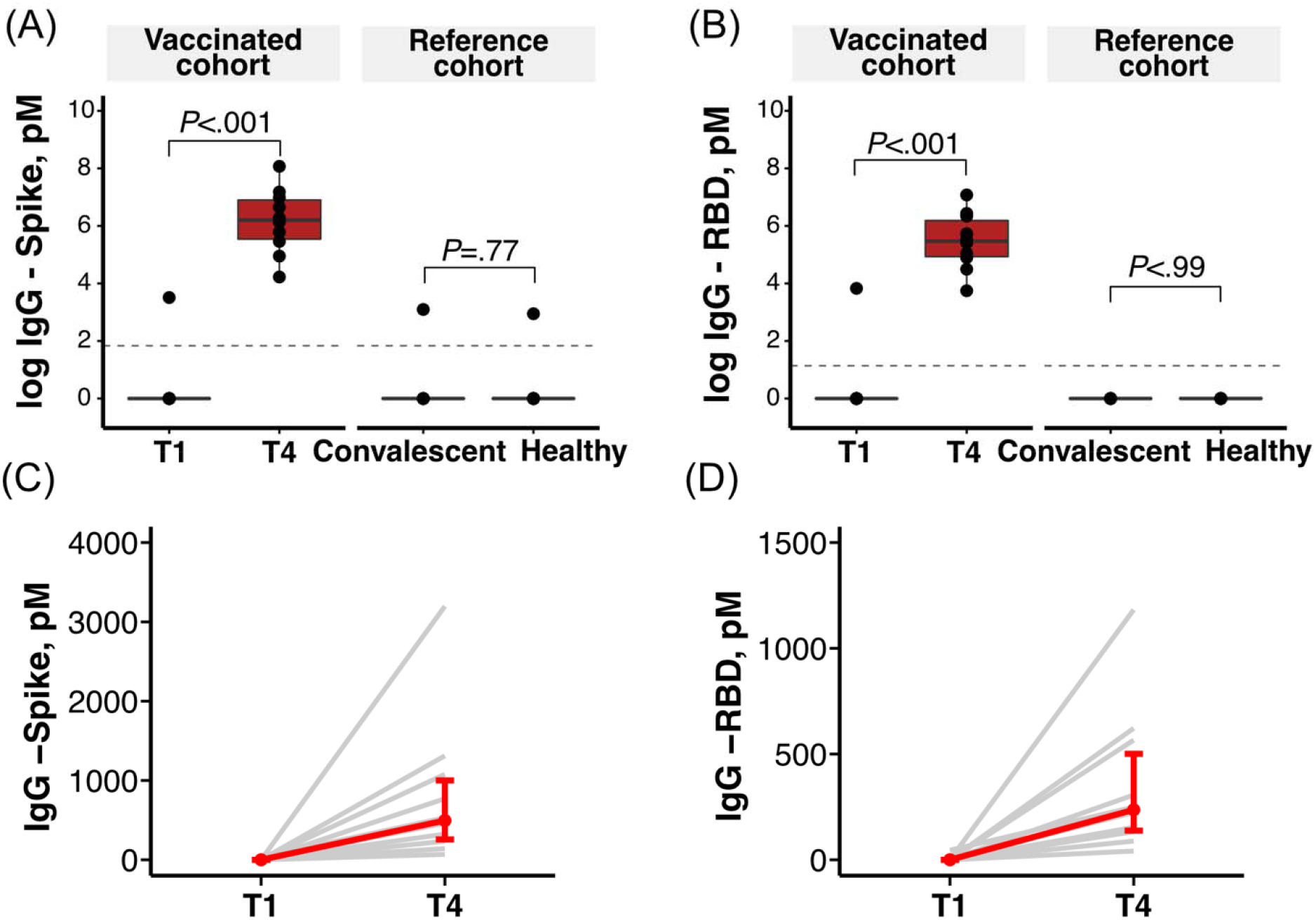
IgG antibodies in human milk samples. (A) & (B): Concentration [pM^a^] of IgG antibodies against spike (A) and RBD (B) in human milk for vaccinated mothers (n = 10) at T1 and T4; convalescent (n = 6) and healthy mothers (n = 9) used as reference cohort. Grey dotted lines reflect the limit of assay detection. Statistics were calculated with a two-tailed Wilcoxon test. (C) & (D): Increase in IgG antibodies against spike (C) and RBD (D) over time. Each line in grey represents data from one individual and the median ± IQR is represented in red. Abbreviations: RBD, receptor-binding domain; IgG, immunoglobulin G; pM, picomolar; IQR, interquartile range; T1, pre-vaccination; T4, 3-7 days after dose two of BNT162b2 vaccine. ^a^ SI conversion factors: To convert concentration from pM to M, multiply values by 10^12^.

Vaccination induced a strong IgA response at T4 (i.e., 3-7 days after dose two of COVID-19 mRNA vaccine). Human milk samples from vaccinated mothers from T4 had medians of 1110 picomolar (pM) (Q1: 672, Q3: 1947; IQR: 1275) of anti-SARS-CoV-2 spike (Figure 1A) and 374 pM (Q1: 240, Q3: 588; IQR: 348) of anti-RBD IgA (Figure 1B), both significantly higher than the concentrations from earlier time points (*P* < .001). Human milk samples evaluated at T4 also exhibited significantly higher levels of IgA compared to the reference samples provided by convalescent mothers who had antenatal COVID-19. IgA was not detected in one mother at T4 (Figure 1C, D; eFigure 2C, D in the Supplement).

As the IgA response from vaccination was observed only at T4, we compared the concentration of IgG targeting spike and RBD in human milk samples collected at T1 and T4. The median concentrations of anti-Spike and anti-RBD IgG at T4 were 495 pM (Q1: 257, Q3: 1001; IQR: 744) and 236 pM (Q1: 138, Q3: 501; IQR: 363), respectively. These levels were significantly higher compared to the concentration at T1 (*P* < .001), and to the convalescent cohort, all of which were negligible (Figure 2A, B; eFigure 3A, B in the Supplement). There was no IgA or IgG detected in healthy unvaccinated lactating women.

**Figure 3.**
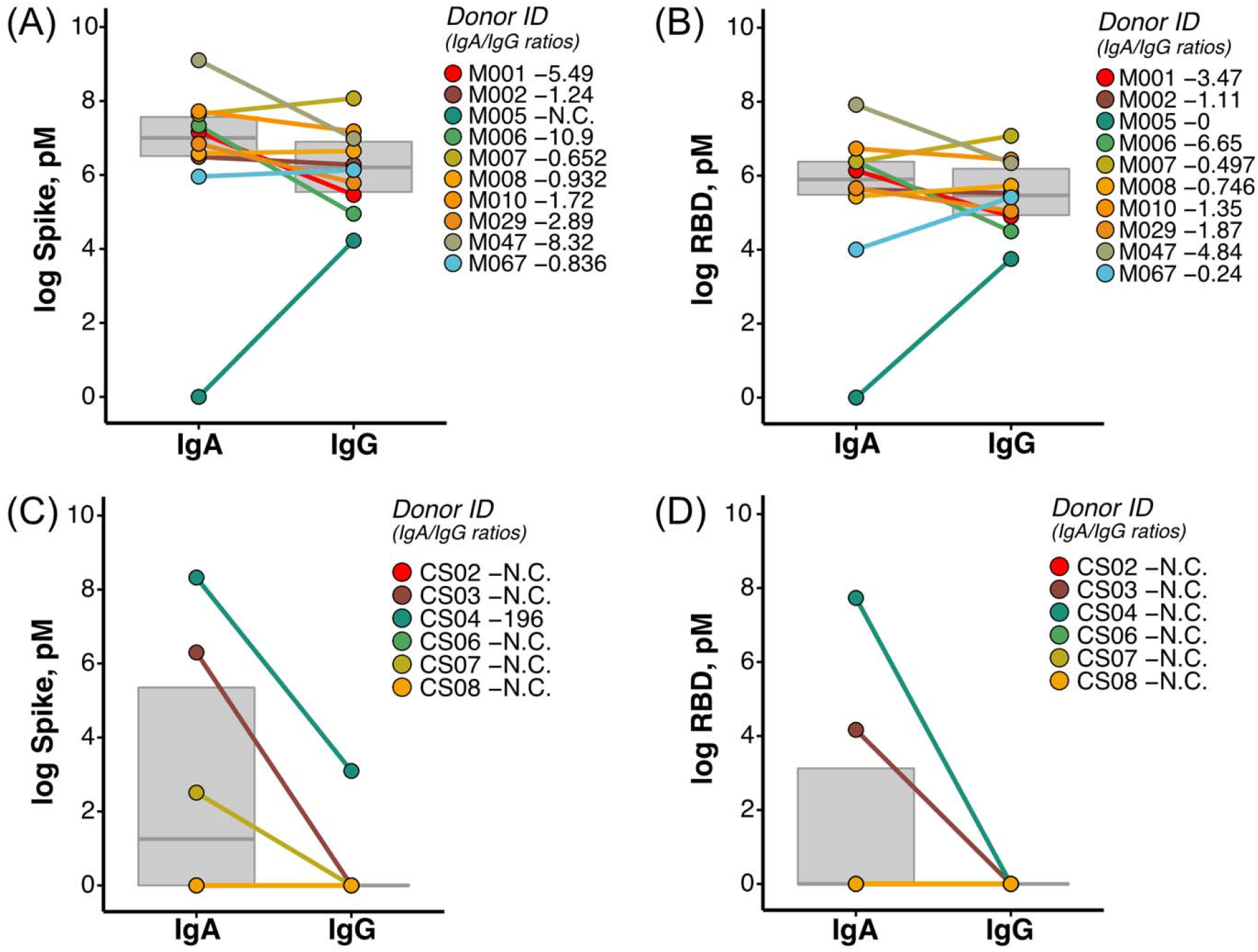
Vaccinated mothers have both IgA and IgG in human milk unlike convalescent mothers. (A) & (B): Concentration of spike (A) and RBD (B) IgA and IgG in vaccinated mothers (n = 10). (C) & (D): Concentration of spike (C) and RBD (D) IgA and IgG in convalescent mothers (n = 6). Legend demonstrates the ratio of IgA and IgG in human milk, whereby the concentration of IgA was divided by the concentration of IgG antibodies. Abbreviations: RBD, receptor-binding domain; IgA, immunoglobulin A; IgG, immunoglobulin G; pM^a^, picomolar; IQR, interquartile range; N.C., not calculated due to absence of either IgA or IgG antibodies. ^a^ SI conversion factors: To convert concentration from pM to M, multiply values by 10^12^.

An increase in IgG in human milk samples were observed in all ten (100%) human milk samples after the second dose of the mRNA vaccine at T4 (Figure 2C, D; eFigure 3C, D in the Supplement).

At T4, both SARS-CoV-2 antigen-specific IgA and IgG antibodies were present in human milk. The ratio of SARS-CoV-2 antigen-specific IgA and IgG was calculated. A ratio of more than 1 would reflect more IgA present than IgG. Conversely, a ratio of less than 1 would reflect more IgG than IgA.

Optical density at 450 nanometer (OD_450_) is a raw assay readout for quantitative ELISA that is highly dependent on factors such as the sensitivity of the test kit. Utilizing this raw assay readout, OD_450_ values of IgG were observed to be above that of IgA in seven (70%) of the vaccinated individuals for both spike (eFigure 4A in the Supplement) and RBD (eFigure 4B in the Supplement).

**Figure 4.**
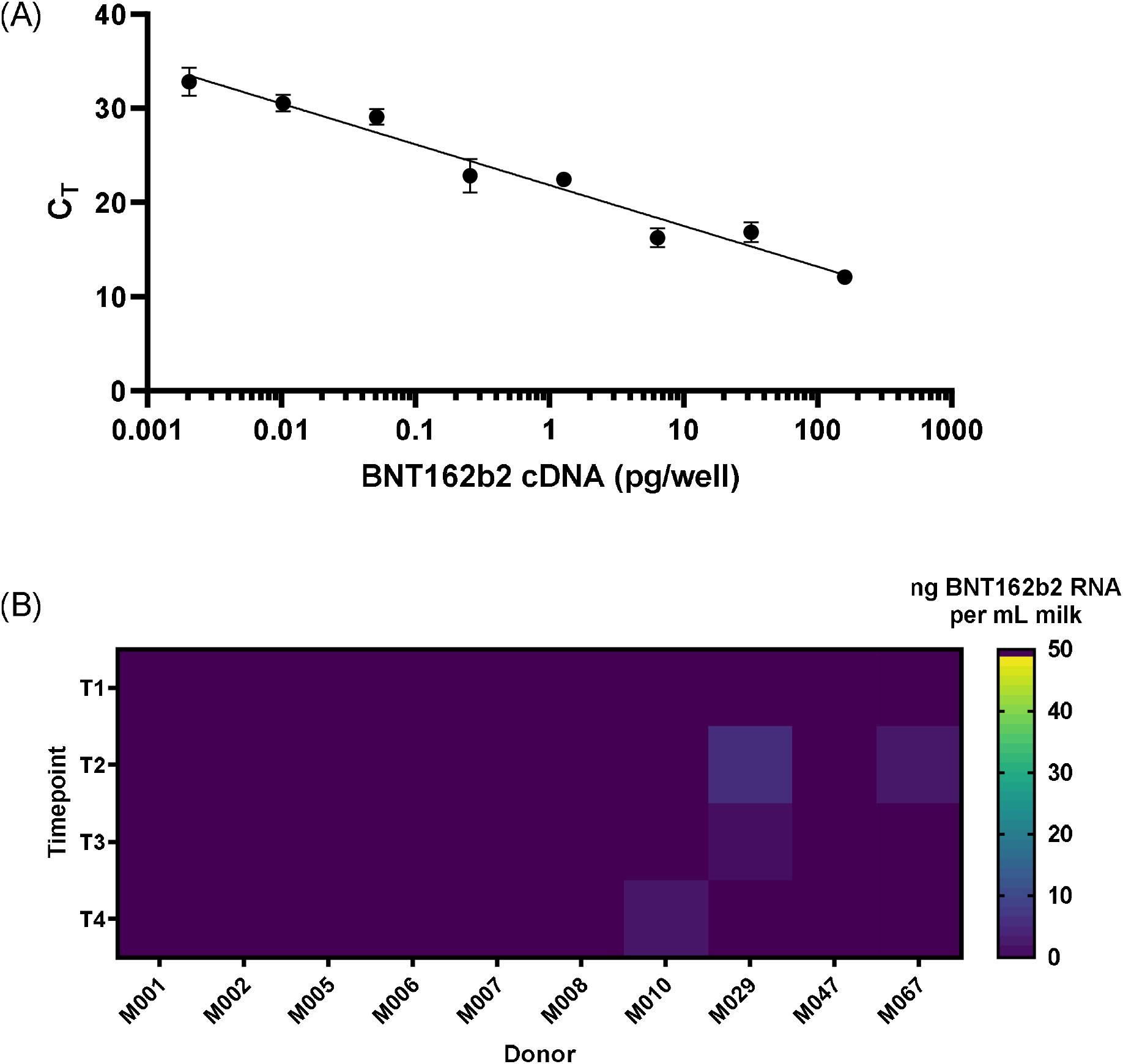
Minimal transfer of BNT162b2 mRNA into human milk. (A) Standard curve of cDNA reverse-transcribed from vaccine derived BNT162b2-spiked human milk was made and used as a positive control. n ≥ 6 technical replicates; means are shown with error bars indicating the standard error of measurement (SEM). (B) Heat map of calculated median BNT162b2 mRNA concentrations in vaccinated mothers (n = 10) at four time points as indicated. Abbreviations: C_T_, cycle threshold; pg^a^, picogram; ng^b^, nanogram; mL^c^, milliliter; T1, pre-vaccination; T2, 1-3 days after dose one of BNT162b2 vaccine; T3, 7-10 days after dose one of BNT162b2 vaccine; T4, 3-7 days after dose two of BNT162b2 vaccine. ^a^ SI conversion factors: To convert concentration from pg to kg, multiply values by 10^15^. ^b^ SI conversion factors: To convert concentration from ng to kg, multiply values by 10^12^. ^b^ SI conversion factors: To convert concentration from mL to L, multiply values by 0.001.

However, in order to perform direct cross-comparison between different antibody isotypes, OD_450_ values were further transformed to picomolar (pM) as the latter quantifies the absolute concentration of antibodies. Here, breaking-down the analysis by individual mothers, we report a higher molar concentration of IgA compared to IgG in six (60%) of the mothers for anti-spike (Figure 3A) and anti-RBD (Figure 3B) antibodies.

Notably, IgG for both spike and RBD were measurable in all ten samples at T4 (i.e., 3-7 days from the dose two) (Figure 3A, B). In contrast, in the convalescent group, spike IgG antibodies were only detected in one woman (Figure 3C, eFigure 4C in the Supplement), and RBD IgG antibodies were not detected in any samples (Figure 3D, eFigure 4D in the Supplement).

### Minimal transfer of BNT162b2 mRNA in milk samples of vaccinated mothers

There was minimal transfer of BNT162b2 mRNA into human milk across all time points. Our assay method could detect sub-picogram levels of BNT162b2 mRNA (Figure 4A). The standard curves constructed with cDNA from spiked milk were similar to that obtained with synthetic DNA constructs (eFigure 5 in the Supplement), indicating robustness of the method. We were only able to observe on rare occasions very low levels of vaccine mRNA in human milk collected within the first week after either dose one or dose two (Figure 4B); 36 out of 40 (90%) samples did not show detectable levels of vaccine mRNA. The highest concentration of BNT162b2 mRNA in the tested samples was 2 ng/mL. This would translate into a hypothetical 0.667% of the original vaccine dose being transferred in 100 mL of human milk given to the infant post vaccination, in the worst-case scenario.

## Discussion

We demonstrated that human milk from lactating mothers who received BNT162b2 vaccines contained SARS-CoV-2-specific antibodies. Within 3-7 days after administering the second dose of BNT162b2 vaccine, nine (90%) individuals produced SARS-CoV-2 IgA, and ten (100%) individuals produced IgG in human milk. The transfer of antibodies via milk may confers local mucosal protection to the breastfed infant. We detected negligible amounts of BNT162b2 mRNA in a minority of human milk samples using a very sensitive assay. Infants in our cohort had no reported adverse events, up to 28 days after ingestion of post-vaccination human milk.

We showed that the sharpest rise in antibody production was after dose two of the BNT162b2 vaccine, with an average of 1882 pM of anti-SARS-CoV-2 spike IgA and 606 pM of anti-RBD IgA being seen in the human milk at 3-7 days after dose two. This is similar to the kinetics found in Baird et al. and Friedman et al., and could either be a function of time after the initial antigenic stimulation of dose one, or an immunological need for dose two.^11,12^ As the humoral immunological response in lactating individuals has been recently shown^18^ to be more dependent on the booster dose of vaccination compared to non-lactating individuals, the latter is highly probable. Thus, whether significant amounts of antigen-specific antibodies can be elicited after administering one-dose regime vaccines, requires confirmation.^18^

Using women convalescent from antenatal COVID-19 as a reference cohort, we were also able to demonstrate that antibody levels immediately after vaccination were significantly higher than those of the convalescent cohort, whose infection was a mean of 5.5 months before the point of human milk collection. This suggests that even for lactating women who have had natural COVID-19, vaccination may be helpful to induce and boost transfer of SARS-CoV-2-specific antibodies in human milk as these protective antibodies wane over time.

Other groups have also found SARS-CoV-2 spike and RBD-specific IgG^11-14^ in human milk after vaccination in addition to IgA. Here, we showed that SARS-CoV-2 spike and RBD-specific IgG are found at significantly higher levels in T4 compared to T1. The presence of vaccine-elicited IgG has been described after intramuscular influenza vaccination,^5^ and is postulated to be related to the intramuscular route of antigenic exposure during vaccination. Compared to a mucosal exposure during natural infection, this would induce increased class-switching to favor IgG as the dominant isotype rather than IgA in human milk.^11^ When comparing OD_450_ raw values for IgA and IgG, a higher OD_450_ for IgG compared to IgA for spike and RBD was detected, suggesting that dominance of the IgG isotype might occur in BNT162b2 vaccinated mothers. However, when converted to picomolar (pM), we show that the majority of vaccinated mothers have increased IgA over IgG, indicating that in human milk, IgA is still the predominant isotype elicited in response to the vaccine.

The role of antigen-specific IgG in human milk is unclear at this time, since IgG does not have a secretory chain and is prone to digestion by the breastfeeding infant; this lends voice to the move to develop intranasal vaccines for respiratory diseases.^19^ However, our study did demonstrate a clear and predominant production of spike-specific IgA, which is resistant to degradation by digestion, which can be expected to confer mucosal immunity to the infant.

In terms of mRNA detection, it is worth noting that our method offers significant advantages over that reported previously.^15^ Phenol-chloroform extraction, the gold standard for RNA extraction, and double-quencher qPCR probes were utilized in our protocol, enhancing the sensitivity of our approach. Consequently, we could detect as low as 2 femtograms (2 × 10^−15^ g) of BNT162b2 cDNA input, which marks an approximately 60-fold increase in sensitivity relative to Golan and colleagues,^15^ where they could not detect vaccine mRNA in human milk up to 48 hours post-vaccination. However, translation and inferred persistence of nanoparticle-delivered mRNA on the scale of days have been previously reported in *in vivo* models.^20^ Thus, we interrogated human milk for presence of mRNA up to one week post vaccination in case of delayed kinetics. In addition, it is important to note that our protocol was designed for specific detection of intact BNT162b2 mRNA, rather than its degraded products, which would be more reflective of whole vaccine components.

Reassuringly, our data suggests that in most cases, vaccine mRNA does not escape into mammary secretions. The few instances where extremely low levels of BNT162b2 mRNA were detected may be due to naturally occurring inter-individual variations in protein adsorption.^19^ This miniscule amount of mRNA is expected to be readily destroyed by enzymes in the infant’s gut, and any accompanying lipid nanoparticles that are excreted into human milk would also be readily digested if ingested orally by the infant.^6,21^

This study has several important strengths. Firstly, we accurately quantified antigen-specific IgA and IgG in human milk. This was referenced against a cohort of lactating women who are convalescent from antenatal COVID-19 and a cohort of unvaccinated lactating women.

Secondly, we used a gold-standard method for detection of vaccine mRNA in human milk, with actual vaccine derived BNT162b2 used as positive control. Lastly, we tracked and reported the clinical status of infants who were fed human milk from vaccinated mothers.

This study has limitations. Firstly, we did not perform any functional assays; however, binding affinity of spike- and RBD-specific antibodies have been correlated with neutralization.^2^ Secondly, the relatively short duration of follow-up period also did not allow for characterization of durability of IgA in human milk after vaccination. Sample collection is ongoing for subsequent studies on antibody durability.

These results lend immunological and clinical evidence to the current recommendation of the ACOG, RCOG and WHO that lactating individuals should continue breastfeeding in an uninterrupted manner after receiving COVID-19 mRNA vaccines.^7,8,22^

## Data Availability

All data referred to in the manuscript are available via correspondence.

## Article Information

### Author Contributions

Drs LWW and YJZ had full access to all of the data in the study and take responsibility for the integrity of the data and the accuracy of the data analysis.

*Concept and design:* JML, YG, MSFN, LYL, ZA, PAM, LWW, YJZ.

*Acquisition, analysis, or interpretation of data:* JML, YG, MSFN, BS, YXN, RG, LWW, YJZ.

*Drafting of the manuscript:* JML, YG, MSFN, LWW, YJZ.

*Critical revision of the manuscript for important intellectual content:* All authors.

*Statistical analysis:* MSFN, YJZ.

*Obtained funding:* JML, ZA, PAM, YJZ.

*Supervision:* LYL, ZA, PAT, PAM.

## Acknowledgements

The authors wish to thank Dr Dimple Rajgor for helping with reviewing, formatting and in submission of the manuscript for publication. We are also deeply grateful to all mothers who donated their time and precious gift of milk to science.

## Online-Only Supplements

### eMethods

Determination of antibody titers in human milk

Sensitive quantitation of BNT162b2 mRNA in human milk

### eTables

**eTable 1:** Demographic and clinical details.

| **Demographic and clinical details** | **Reference cohort (n = 6)** | **Control cohort (n = 9)** |
| --- | --- | --- |
| Age, mean [standard deviation, SD], years | 30.8 [4.1] | 32.1 [4.1] |
| Ethnicity | 1 Chinese<br>2 Malay<br>1 Indian<br>2 Caucasian | 6 Chinese<br>1 Malay<br>2 Indian |
| Pre-existing comorbidities | 5 None<br>1 Pre-existing hepatitis C | 8 None<br>1 Pre-existing hepatitis C |
| Antenatal conditions | 6 None | 9 None |
| Antenatal ultrasound abnormalities | 6 No abnormalities | 2 Intrauterine growth restriction |
| Gestational age at delivery, mean [SD], weeks | 40.0 [0.8] | 39.2 [1.2] |
| Mode of delivery | 6 Vaginal | 6 Vaginal<br>3 Caesarean section |
| Gender of infants | 5 Female | 6 Female |
| Infant birth weight, mean [SD], kg | 3.59 [0.24] | 3.11 [0.61] |
| Infant birth length, mean [SD], cm<sup>a</sup> | 50.8 [1.6] | 50.5 [2.2] |
| Infant birth occipital frontal circumference, mean [SD], cm<sup>a</sup> | 34.6 [0.9] | 22.3 [1.2] |
| Any immediate postnatal complications | 5 None<br>1 Transient tachypnoea of the newborn | 8 None<br>1 Exaggerated physiological jaundice |
| Type of feeds | 4 Exclusive breastfeeding<br>2 Mixed feeding | 6 Exclusive breastfeeding<br>3 Mixed feeding |
| <sup>a</sup> SI conversion factors: To convert length from cm to m, multiply values by 0.01. |  |  |

**eTable 2:**
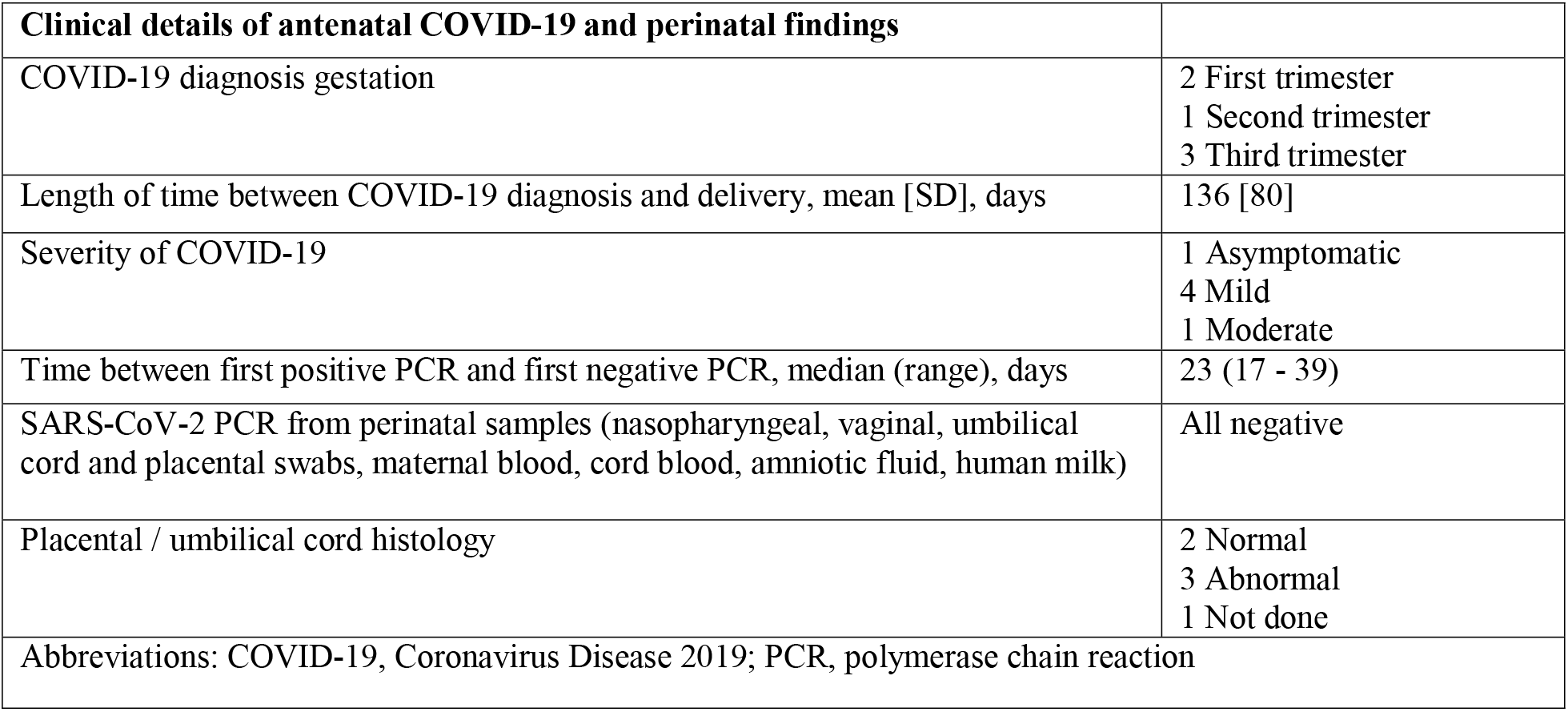
Clinical details of antenatal COVID-19 and perinatal findings.

| **Clinical details of antenatal COVID-19 and perinatal findings** |  |
| --- | --- |
| COVID-19 diagnosis gestation | 2 First trimester<br>1 Second trimester<br>3 Third trimester |
| Length of time between COVID-19 diagnosis and delivery, mean [SD], days | 136 [80] |
| Severity of COVID-19 | 1 Asymptomatic<br>4 Mild<br>1 Moderate |
| Time between first positive PCR and first negative PCR, median (range), days | 23 (17 - 39) |
| SARS-CoV-2 PCR from perinatal samples (nasopharyngeal, vaginal, umbilical cord and placental swabs, maternal blood, cord blood, amniotic fluid, human milk) | All negative |
| Placental / umbilical cord histology | 2 Normal<br>3 Abnormal<br>1 Not done |
| Abbreviations: COVID-19, Coronavirus Disease 2019; PCR, polymerase chain reaction |  |

### eFigures

**eFigure 1.**
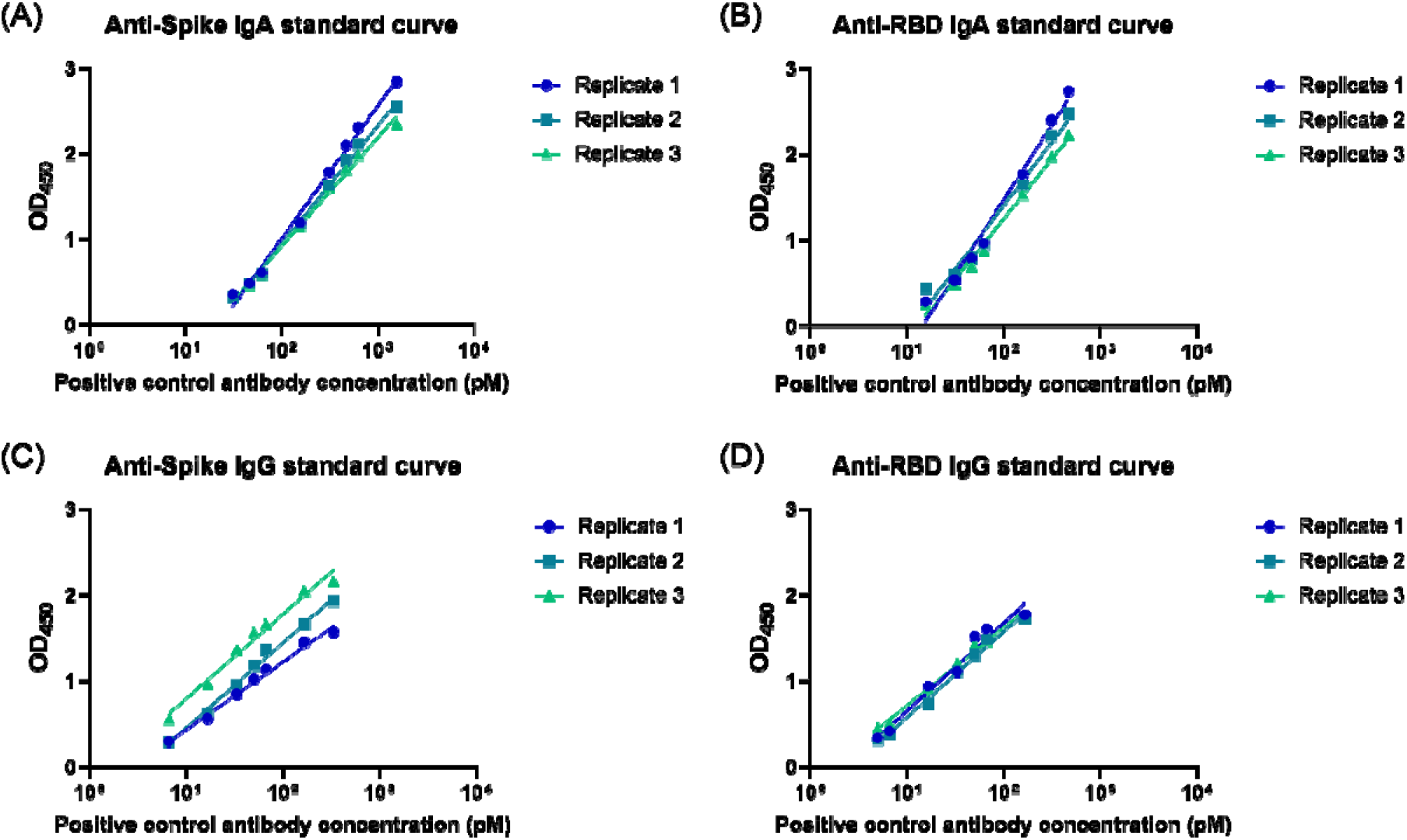
Standard curves used in quantitative ELISA. A human monoclonal antibody binding to both full spike and RBD of SARS-CoV-2 was recombinantly expressed as human IgA1 and IgG1. Concentrations of (A) anti-spike IgA, (B) anti-RBD IgA, (C) anti-spike IgG, and (D) anti-RBD IgG antibodies in human milk samples were estimated from interpolation of the standard curves constructed using known concentrations of this antibody (in pM^a^). Abbreviations: RBD, receptor-binding domain; IgA1, immunoglobulin A1; IgG1, immunoglobulin G1; pM, picomolar; OD_450_, optical density measured at a wavelength of 450 nanometer. ^a^ SI conversion factors: To convert concentration from pM to M, multiply values by 10^12^.

**eFigure 2.**
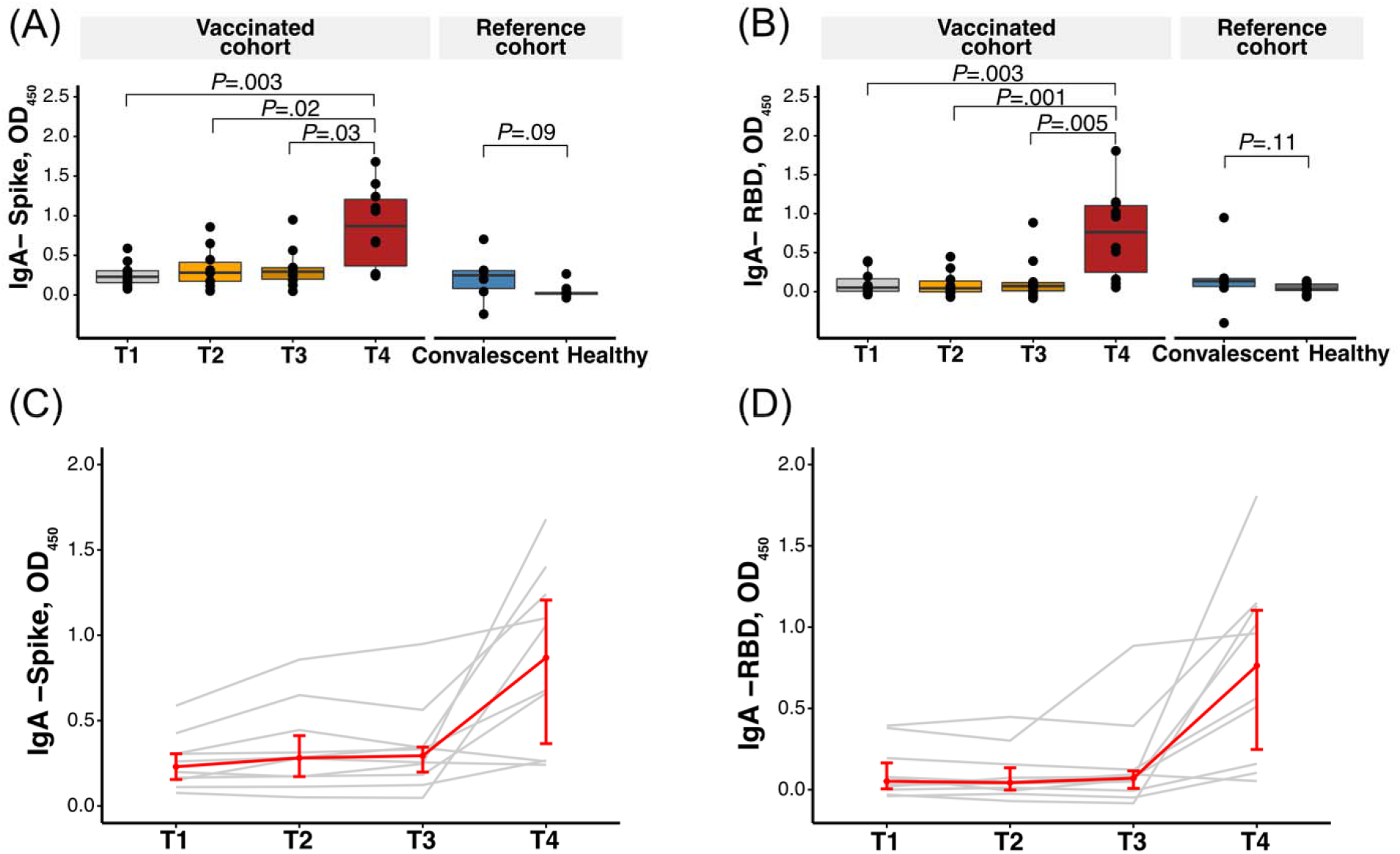
SARS-CoV-2 antigen-specific IgA levels in human milk samples. OD_450_ values were measured for (A) anti-spike and (B) anti-RBD IgA antibodies in 10-fold diluted human milk samples collected from vaccinated mothers (n = 10) at four time points pre- and post-vaccination. Statistics were calculated with Kruskal-Wallis test with Dunn’s post-test. The same results are shown as line plots in (C) and (D) for visualization of antibody response from each individual. Each line in grey represents data from one individual and the median ± IQR is represented in red. Abbreviations: OD_450_, optical density measured at a wavelength of 450 nanometer; RBD, receptor-binding domain; IgA, immunoglobulin A; IQR, interquartile range; T1, pre-vaccination; T2, 1-3 days after dose one of BNT162b2 vaccine; T3, 7-10 days after dose one of BNT162b2 vaccine; T4, 3-7 days after dose two of BNT162b2 vaccine.

**eFigure 3.**
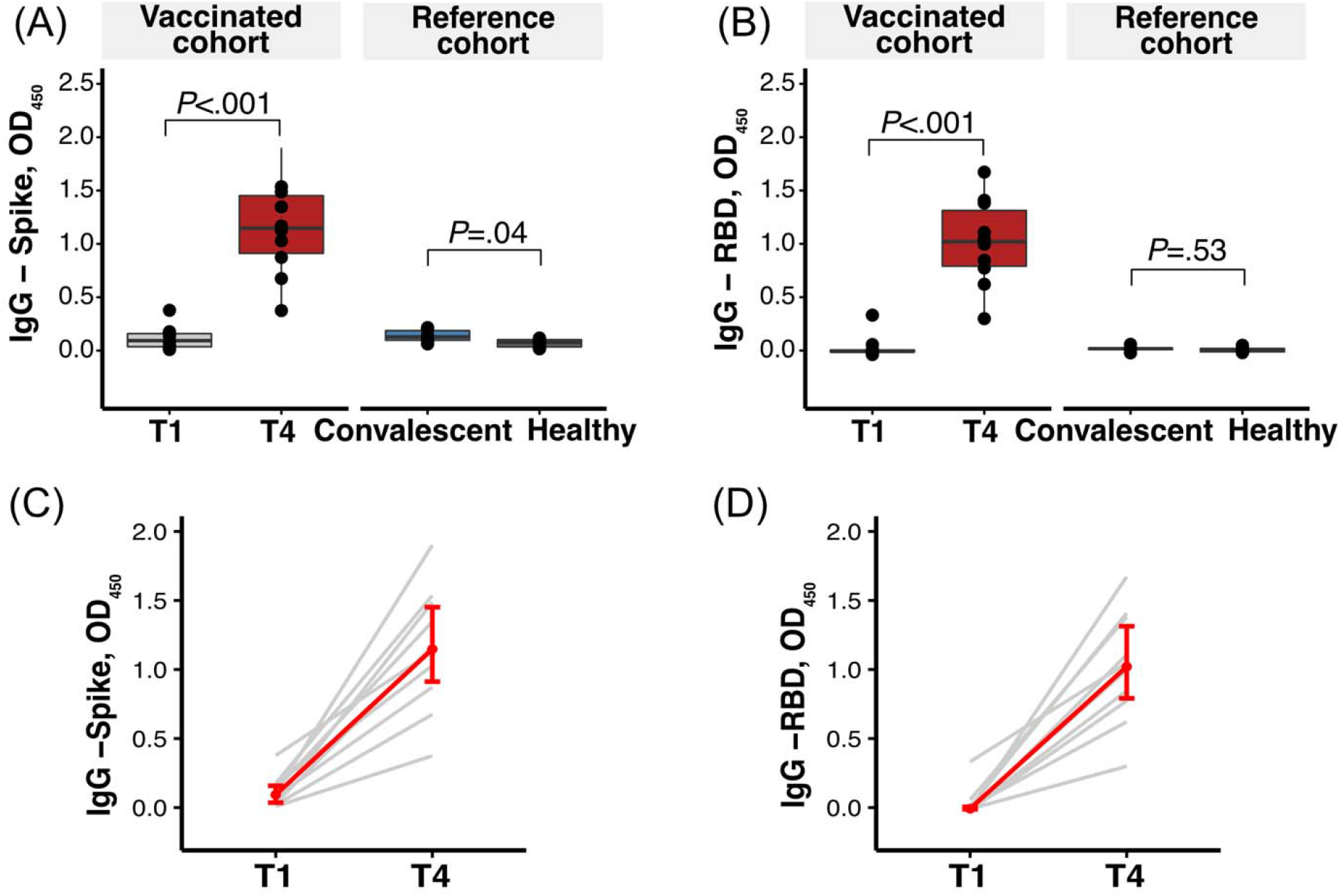
SARS-CoV-2 antigen-specific IgG levels in human milk samples. OD_450_ values were measured for (A) anti-spike and (B) anti-RBD IgG antibodies in 10-fold diluted human milk samples collected from vaccinated mothers (n = 10) at two time points pre-(T1) and post-second dose vaccination (T4). The same results are shown as line plots in (C) and (D) for visualization of antibody response from each individual. Each line in grey represents data from one individual and the median ± IQR is represented in red. Abbreviations: OD_450_, optical density measured at a wavelength of 450 nanometer; RBD, receptor-binding domain; IgG, immunoglobulin G; IQR, interquartile range; T1, pre-vaccination; T4, 3-7 days after dose two of BNT162b2 vaccine.

**eFigure 4.**
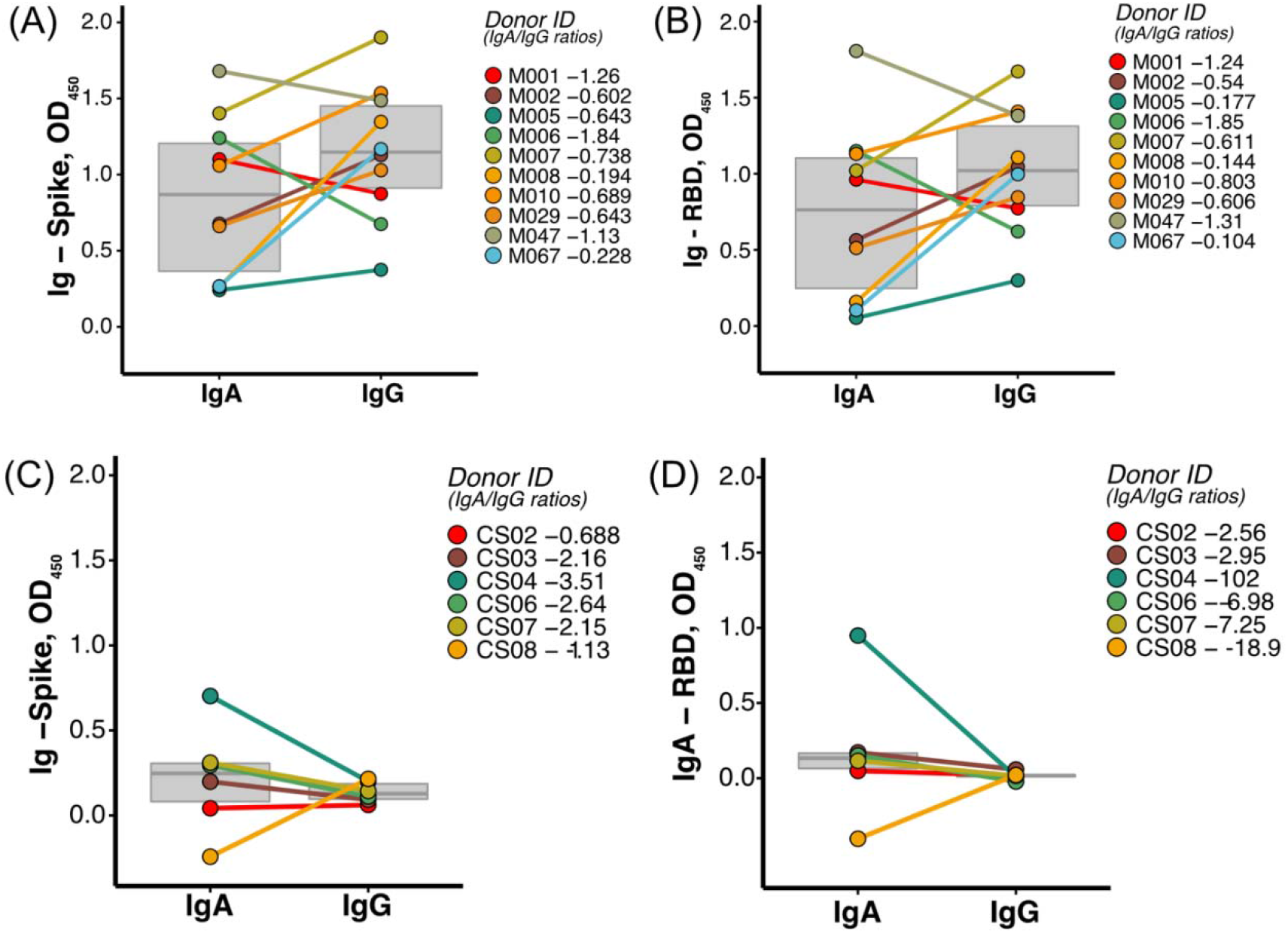
Cross-comparison of SARS-CoV-2 antigen-specific IgA and IgG levels in human milk samples. (A,B) OD_450_ values were measured for the vaccination cohort and show (A) anti-spike and (B) anti-RBD antibodies in 10-fold diluted human milk samples collected from vaccinated mothers (n = 10) at 3-7 days after the second vaccination dose (T4). (C,D) OD_450_ values were measured for the reference cohort and show (C) anti-spike and (D) anti-RBD antibodies in 10-fold diluted human milk samples for cases (n = 6) at one month post-partum. Abbreviations: OD_450_, optical density measured at a wavelength of 450 nanometer; RBD, receptor-binding domain; IgA, immunoglobulin A; IgG, immunoglobulin G; IQR, interquartile range.

**eFigure 5.**
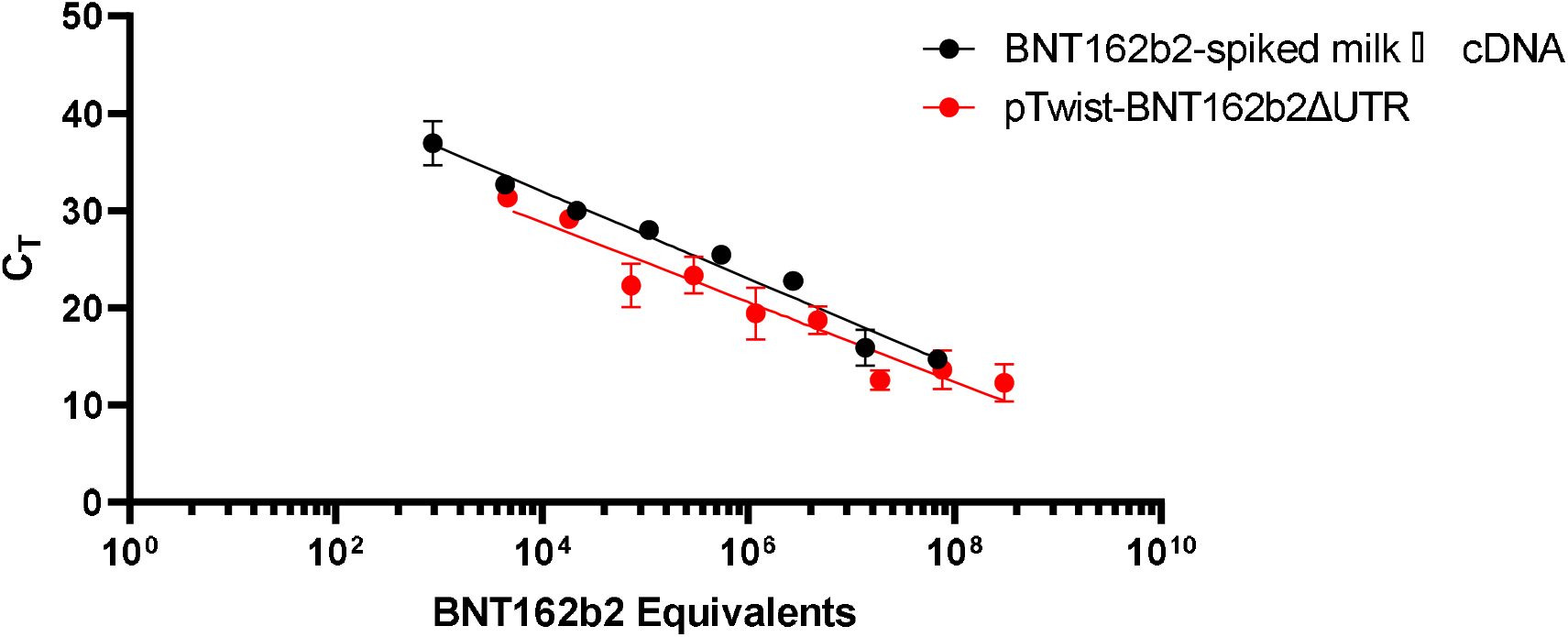
Standard curves using cDNA generated from BNT162b2-spiked human milk versus pTwist-BNT162b2ΔUTR plasmid. One BNT162b2 equivalent is defined as a single-stranded polynucleotide molecule that contains a linear sequence that binds either the forward or reverse primer. Prism analysis of the two regression lines revealed that their slopes/gradients were not significantly different (*P* = .42). Data represented as the mean ± SEM, n ≥ 3 technical replicates. Abbreviations: cDNA, complementary DNA; C_T_, cycle threshold.

### Detailed methods

#### Determination of antibody titers in human milk

IgA and IgG against SARS-CoV-2 antigens including the whole spike and RBD protein were titrated using quantitative ELISA. 96-well flat-bottom maxi-binding immunoplate (SPL Life Sciences, #32296) were coated with 100 ng of SARS-CoV-2 whole spike protein or 200ng of RBD protein at 4 °C overnight. After three washes in Phosphate Buffer Saline (PBS), 350 µL of blocking buffer [4% skim milk in PBS with 0.05% Tween 20 (PBST)] was added to each well. After incubation for 1.5 hours, the plate was washed three times with PBST. 100 µL of 10-fold diluted human milk samples were added to each well for 1-hour incubation. Plate was then washed three times with PBST followed by 1-hour incubation in the dark with 100 µL of 5000-fold diluted goat anti-human IgG-HRP (Invitrogen, #31413), or 5000-fold diluted F(ab’)2 anti-human IgA-HRP (Invitrogen, #A24458). Plate was washed three times in PBST and incubated for 3 minutes with 1-Step Ultra TMB-ELISA (Thermo Scientific, #34029), 100 µL per well. Reaction was stopped with 100 µL of 1 M H_2_SO_4_ and OD_450_ was measured using a microplate reader (Tecan Sunrise). OD_450_ was calculated by subtracting the background signal from sample binding to the blocking buffer. A recombinant human monoclonal antibody targeting RBD of SARS-CoV-2 was engineered to both human IgG1 and human IgA1. Standard curves for whole spike and RBD were constructed by testing known concentrations (3.5 pM - 3.5 nM) of this antibody alongside the samples. Linear region of the standard curve was used to quantify the IgG/IgA antibody concentration in human milk samples via interpolation. Experiments were performed at least three times.

#### Sensitive quantitation of BNT162b2 mRNA in human milk

Total RNA from human milk was extracted with TRIzol LS reagent (Invitrogen) according to manufacturer’s instructions. Briefly, 250 μL of whole human milk was RNA-extracted with 750 μL of Trizol LS and 200 μL of chloroform. The entire upper aqueous fraction (∼500 μL) was isopropanol-precipitated, washed with 70% ethanol and air-dried before dissolution in RNAse-free water. The entire volume of RNA was used as input for SuperScript IV (Invitrogen) reverse transcription performed as per manufacturer’s instructions and diluted 50-fold with RNAse-free water to a final volume of 1 mL prior to storage at −20□C or assay.

For standard curve construction, we collected BNT162b2 from actual vaccine discards. Five-fold serial dilutions of reconstituted BNT162b2 vaccine were spiked into healthy, SARS-CoV-2 negative human milk and RNA-extracted in parallel with vaccinee samples. These served as a positive control.

Taqman-based detection of BNT162b2 mRNA was performed using PrimeQuest-designed primer and probe sets (Integrated DNA Technologies, IDT). The primer and probe sequences are as follows: AGCCTACACCAACAGCTTTAC (forward primer), TGAAGAAAGGCAGGAACAGG (reverse primer) and /56-FAM/CGACAAGGT/ZEN/GTTCAGATCCAGCGT/3IABkFQ/ (probe). Primers were used at 250 nM while probe was used at 150 nM. An input volume of 1 μL was used for each diluted cDNA sample. Standard cycling conditions used were as per recommendation by IDT.

A plasmid construct containing BNT162b2 cDNA without the 5’ and 3’ UTRs was synthesized (Twist Bioscience) and is herein referred to as pTwist-BNT162b2ΔUTR. Briefly, the BNT162b2 open reading frame is 3825 bases long and consists of human codon-optimized SARS-CoV-2 spike glycoprotein containing the K986P and V987P mutations^1^

Standard curve construction was performed in a separate experiment using pTwist-BNT162b2ΔUTR and cDNA generated from BNT162b2-spiked human milk with the aim of determining whether their performances in the quantitative PCR assay were comparable.

